# Lower risks of sodium glucose cotransporter 2 (SGLT2) inhibitors compared to dipeptidyl peptidase-4 (DPP4) inhibitors for new-onset hip fracture risks in patients with type-2 diabetes: A propensity score-matched study with competing risk analysis

**DOI:** 10.1101/2023.02.13.23285846

**Authors:** Oscar Hou In Chou, Jiandong Zhou, Danish Iltaf Satti, Jonathan V Mui, Karen Lin, Sharen Lee, Wing Tak Wong, Abraham Ka Chung Wai, Tong Liu, Bernard Man Yung Cheung, Fengshi Jing, Gary Tse

**Author notes:** Correspondence to: Bernard Man Yung Cheung MB BChir PhD FRCP, Department of Medicine, LKS Faculty of Medicine, University of Hong Kong, Hong Kong, China, Fengshi Jing, PhD, Institute for Artificial Intelligence, Guangdong Second Provincial General Hospital, Guangzhou, China, The University of North Carolina at Chapel Hill Project-China, Guangzhou, China., Gary Tse, MD, DM, PhD, Tianjin Institute of Cardiology, The Second Hospital of Tianjin Medical University Tianjin 300211, China, Kent and Medway Medical School, Canterbury Christ Church University and University of Kent, Canterbury, United Kingdom, School of Nursing and Health Studies, Metropolitan University, Hong Kong, China. Joint first authors (Oscar Hou In Chou, Jiandong Zhou).

## Abstract

**Purpose:** This study aimed to compare the effects of sodium glucose cotransporter 2 inhibitors (SGLT2I) and dipeptidyl peptidase-4 inhibitors (DPP4I) on new-onset hip fractures.

**Methods:** This was a retrospective population-based cohort study including type-2 diabetes mellitus patients treated with either SGLT2I or DPP4I between January 1^st^ 2015 and December 31^st^ 2020 in Hong Kong. The primary outcome was new-onset hip fracture and the secondary outcome was all-cause mortality. Propensity score matching (1:1 ratio) using the nearest neighbour search was performed. Univariable and multivariable Cox regression were applied to identify significant predictors. Competing risks models and multiple approaches using the propensity score were performed.

**Results:** This cohort included 56393 patients with type-2 diabetes mellitus (median age: 62.1 years old [interquantile range, IQR]: 54.2-71.1; 57.45% males), of which 20432 patients ([incidence rate, IR]: 36.23%) used SGLT2I and 35961 patients (IR: 63.77%) used DPP4I. After the 1:1 propensity score matching, 449 (IR: 1.09%) patients had hip fractures, and 2012 patients (IR: 4.92%) died. SGLT2I was associated with significantly lower risks of hip fractures after adjusting for the demographics, past comorbidities, non-SGLT2I/DPP4I medications and laboratory results (hazard ratio: 0.55; 95% confidence interval: 0.42-0.89; P=0.0036). The results were consistent in the competing risk models and the different propensity matching approaches.

**Conclusions:** SGLT2I was associated with lower risks of new-onset hip fractures after propensity score matching and adjustments.

**Summary:** This study compared the risks of hip fractures in between users of sodium glucose cotransporter 2 inhibitors (SGLT2I) and dipeptidyl peptidase-4 inhibitors in type-2 diabetes mellitus. After propensity score matching, SGLT2I was associated with lower risks of hip fractures adjusting for confounders (hazard ratio: 0.55; 95% confidence interval: 0.42-0.89; P=0.0036).

## Introduction

Type 2 diabetes mellitus (T2DM) is an important component of metabolic syndrome that is becoming increasingly prevalent worldwide. It was forecasted that by 2030, 7079 individuals per 100,000 of the population would be affected by T2DM.^1^ T2DM increases the risks of developing complications such as nephropathy, coronary heart disease, and cerebrovascular disease.^2^ T2DM is associated with higher risks of bone fragility and bone fracture via mechanisms such as increasing the amount of advanced glycation end products (AGEs).^3, 4^ A recent study showed that T2DM is associated with 70% higher risks of hip fracture. Furthermore, the daily living and mobility of diabetic patients are heavily compromised following a hip fracture.^5^ Given the disease burdens of T2DM, novel classes of drugs such as sodium-glucose cotransporters-2 inhibitors (SGLT2I) and dipeptidyl peptidase-4 inhibitors (DPP4I) were introduced to control T2DM.

DPP4I reduces the blood glucose level via increasing the level of glucagon-like peptide-1 (GLP-1). DPP4I was described to reduce the risks of fracture via both glucose-dependent and glucose-independent means.^6^ Previously, it was demonstrated that DPP4I usage significantly reduces the risk of all-cause fractures among T2DM patients after 5 years of follow-up.^7^ Moreover, DPP4I was not associated with higher risks of fracture compared to sulphonylurea and insulin.^8^ Furthermore, GLP-1 was reported to enhance bone formation, inhibit osteoclastic activity, and increase bone quality.^9, 10^ SGLT2I blocks glucose reabsorption at the S1 segment of the proximal convoluted tubules of the kidney to reduce the blood glucose level. In a randomized clinical trial, SGLT2I was demonstrated to be associated with higher risks of fracture compared to those taking placebo.^11^ Canagliflozin, for instance, was reported to decrease the bone mineral density and thus, associated with increased risks of fracture.^12^ However, a meta-analysis showed that SGLT2I was not associated with increased risks of bone fracture compared to the placebo. However, the conclusion was limited by the wide confidence interval.^13^ Moreover, a study demonstrated that the risk of fractures were not different between SGLT2I and DPP4 among elderly patients.^14, 15^

To the best of our knowledge, there has yet to be a study that compares the effects of SGLT2I and DPP4I on hip fracture directly. Therefore, the present study aimed to compare the risk of hip fracture between SGLT2 and DPP4I patients among T2DM patients in Hong Kong.

## Methods

### Study design and population

This was a retrospective, territory-wide cohort study of T2DM patients treated with SGLT2I or DPP4I between January 1^st^, 2015 and December 31^st^, 2020 in Hong Kong. Patients were followed up until December 31^st^, 2020 or until death. This study was approved by The Joint Chinese University of Hong Kong–New Territories East Cluster Clinical Research Ethics Committee. Patients were excluded based on the following criteria: 1) taking both DPP4I and SGLT2I, or switched between the two drug classes; 2) without complete demographic or mortality data; 3) history of human immunodeficiency virus infection; 4) pregnancy; 5) younger than 18 years old at the start of the study; 6) died within 30 days after initial drug exposure; 7) prior diagnosis of osteopenia, osteoporosis, hip fractures, accidental falls, bone tumor, rheumatoid arthritis or other inflammatory polyarthropathies, osteoarthritis and allied disorders or other bone diseases. The patients were identified from the Clinical Data Analysis and Reporting System (CDARS), a territory-wide database that centralizes patient information from individual local hospitals to establish comprehensive medical data, including clinical characteristics, disease diagnosis, laboratory results, and drug treatment details. The system has been used by both our team and other teams in Hong Kong to conduct comparative studies between SGLT2i and DPP4I use ^16, 17^ and hip fractures in other cohorts.^18, 19^

Patients’ demographics including gender and age of initial drug use (baseline), clinical and biochemical data were extracted for the present study. Prior comorbidities were extracted using the *International Classification of Diseases Ninth Edition* (ICD-9) codes (**Supplementary Table 1**). Charlson’s standard comorbidity index was also calculated. Both cardiovascular medications and anti-diabetic agents were also extracted. The baseline laboratory examinations, including the complete blood count, renal and liver biochemical tests, and the lipid and glucose profiles were extracted. The renal function was calculated using the abbreviated modification of diet in renal disease (MDRD) formula.^20^ Mortality was recorded using the *International Classification of Diseases Tenth Edition* (ICD-10) coding.

Standard deviation (SD) was calculated for glycaemic and lipid profile parameters if there were at least three measurements for each patient since initial drug exposure to SGLT2I or DPP4I. Different variability measures, including standard deviation (SD), SD/initial test, coefficient of variation (CV), and variability independent of mean, were also calculated (**Supplementary Table 2**).

### Adverse outcomes and statistical analysis

The primary outcome was new-onset hip fracture and the secondary outcome was all-cause mortality. Mortality data were obtained from the Hong Kong Death Registry, a population-based official government registry with the registered death records of all Hong Kong citizens linked to CDARS. Hip fractures that were recorded with a traumatic event (ICD-9-CM E800-E848) were regarded as censoring events and were not included as outcome events in order to exclude possible cases of traumatic fractures. Only a maximum of one hip fracture (initial hip fracture) per patient was included, and recurrent hip fracture was not considered in the present study. The endpoint date of interest for eligible patients was the event presentation date. The endpoint for those without primary outcome presentation was the mortality date or the endpoint of the study (December 31^st^, 2020).

Descriptive statistics were used to summarize baseline clinical and biochemical characteristics of patients with SGLT2I and DPP4I use. For baseline clinical characteristics, the continuous variables were presented as mean (95% confidence interval [CI]/standard deviation [SD])) and the categorical variables were presented as total numbers (percentage). Continuous variables were compared using the two-tailed Mann-Whitney U test, whilst the two-tailed Chi-square test with Yates’ correction was used to test 2×2 contingency data. Propensity score matching with 1:1 ratio for SGLT2I use v.s. DPP4I use based on demographics, Charlson comorbidity index, prior comorbidities, non-SGLT2I/ DPP4I medications were performed using the nearest neighbor search strategy. We used Stata software (Version 16.0) to conduct the propensity score matching procedures.

Baseline characteristics between patients with SGLT2I and DPP4I use before and after matching were compared using the standardized mean difference (SMD), with SMD<0.20 regarded as well-balanced between two groups. Proportional Cox regression models were used to identify significant risk predictors of adverse study outcomes. Cause-specific and subdistribution hazard models were conducted to consider possible competing risks. Multiple propensity adjustment approaches were used, including propensity score stratification,^21^ propensity score matching with inverse probability of treatment weighting ^22^ and propensity score matching with stable inverse probability weighting.^23^ The hazard ratio (HR), 95% CI and P-value were reported. Statistical significance is defined as P-value < 0.05. All statistical analyses were performed with RStudio software (Version: 1.1.456) and Python (Version: 3.6).

## Results

### Basic characteristics

This was a retrospective, territory-wide cohort study of 76147 patients with T2DM treated with SGLT2I/DPP4I between January 1^st^, 2015 and December 31^st^, 2020 in Hong Kong. Patients during the aforementioned period were enrolled and followed up until December 31^st^, 2020 or until death. Patients with both DPP4I and SGLT2I use (N=12858), without complete demographics (N=17), without mortality data (N=13), with human immunodeficiency virus infection (N=5), with pregnancy (N=9), less than 18 years old (N=135), died within 30 days at initial drug exposure (N=231), with a prior diagnosis of osteopenia (N=7), osteoporosis (N=19), hip fractures (N=1813), osteoarthritis and allied disorders (N=21), or other bone diseases (N=4626) were excluded **(Figure 1)**.

**Fig 1.**
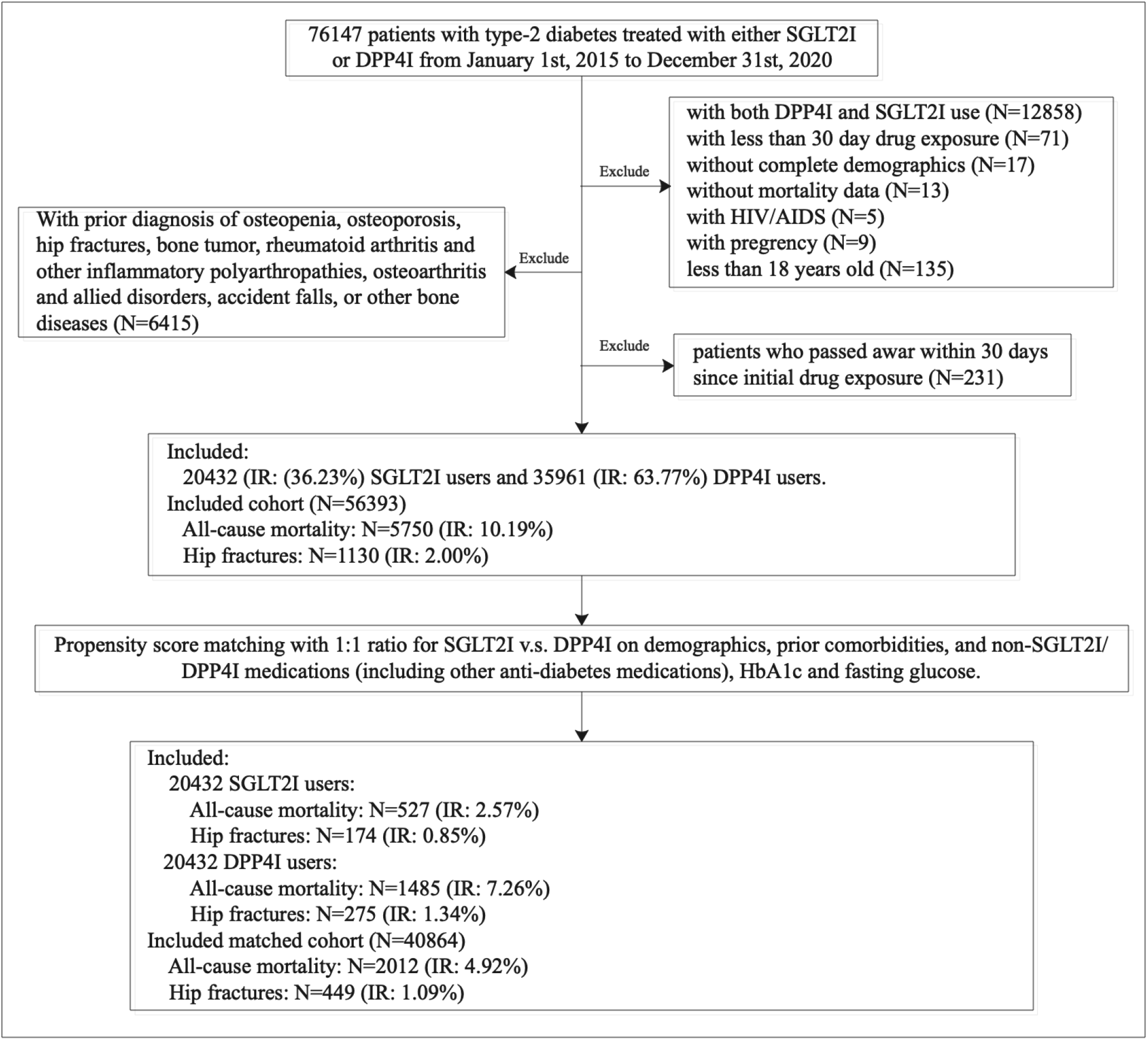
Study Flow and Patient Selection. IR: Incidence rate; SGLT2I: Sodium-glucose cotransporter-2 inhibitors; DPP4I: Dipeptidyl peptidase-4 inhibitors.

After exclusion, this study included a total 56393 patients with T2DM (median age: 62.1 years old [interquantile range, IQR]: 54.2-71.1; 57.45% males). The median follow-up duration was 5.56 (IQR: 5.24-5.8) years since the initial drug exposure. 20432 patients ([incidence rate, IR]: 36.23%) used SGLT2I and 35961 patients (IR: 63.77%) used DPP4I. The propensity score matching comparisons before and after 1:1 matching demonstrated that there were no significant confounding characteristics after matching, and the proportional hazard assumption was fulfilled **(Supplementary Figure 1)**. After 1:1 propensity score matching, 2012 patients (IR: 4.92%) died and 449 (IR: 1.09%) patients developed hip fractures. The characteristics of SGLT2 and DPP4 users are shown in **Table 1**.

**Table 1.**
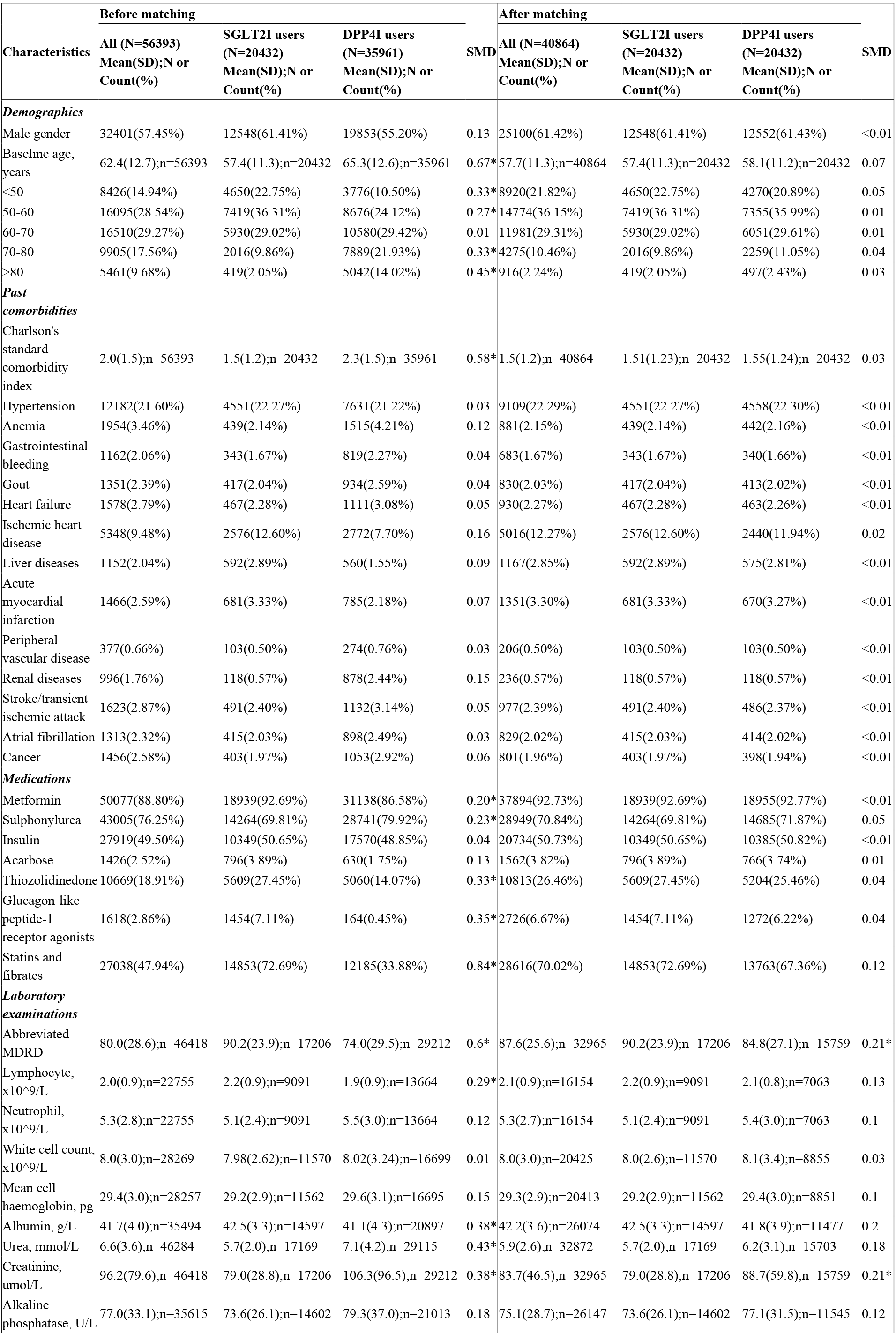
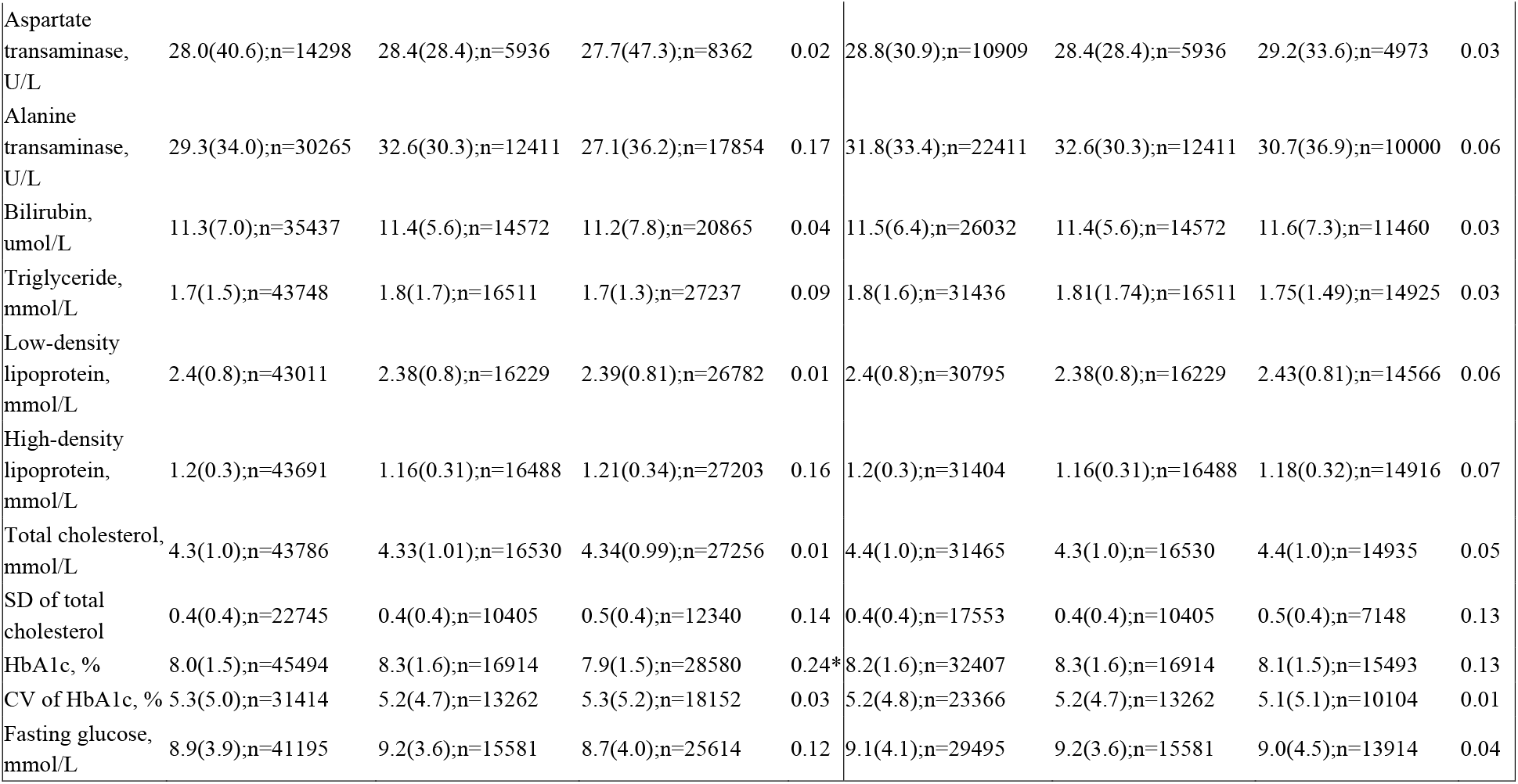
Baseline and clinical characteristics of patients with SGLT2I users v.s. DPP4I users before and after propensity score matching (1:1). * for SMD≥0.2; SD: standard deviation; SGLT2I: sodium glucose cotransporter-2 inhibitor; DPP4I: dipeptidyl peptidase-4 inhibitor; CV: coefficient of variation.

### Significant predictors of the study outcomes

Univariable Cox regression identified the significant risk factors for hip fractures and mortality before and after propensity score matching (1:1) **(Table 2 and Supplementary Table 3)**. In the matched cohort, SGLT2I was associated with lower risks of new-onset hip fracture (HR: 0.62; 95% C: 0.51-0.75; P <0.0001). The cumulative incidence curves for hip fractures and all-cause mortality stratified by SGLT2I versus DPP4I demonstrated that SGLT2I was associated with lower cumulative hazard for hip fractures and all-cause mortality **(Figure 2)**. In the multivariable cox models, SGLT2I was associated with lower risks of new-onset hip fracture (HR: 0.55; 95% CI: 0.42-0.89; P=0.0036) and all cause mortality (HR: 0.35; 95% CI: 0.31-0.55; P<0.0001) after adjusting for the demographics, comorbidities, non-SGLT2I/DPP4I medications, and laboratory results. In the subgroup analysis, patients >65 years old had the highest cumulative hazard for developing a hip fracture among both SGLT2I and DPP4I users **(Supplementary Figure 2)** and SGLT2I users had a lower cumulative hazard for developing hip fracture regardless of age and gender (**Supplementary Figure 3)**.

**Table 2.**
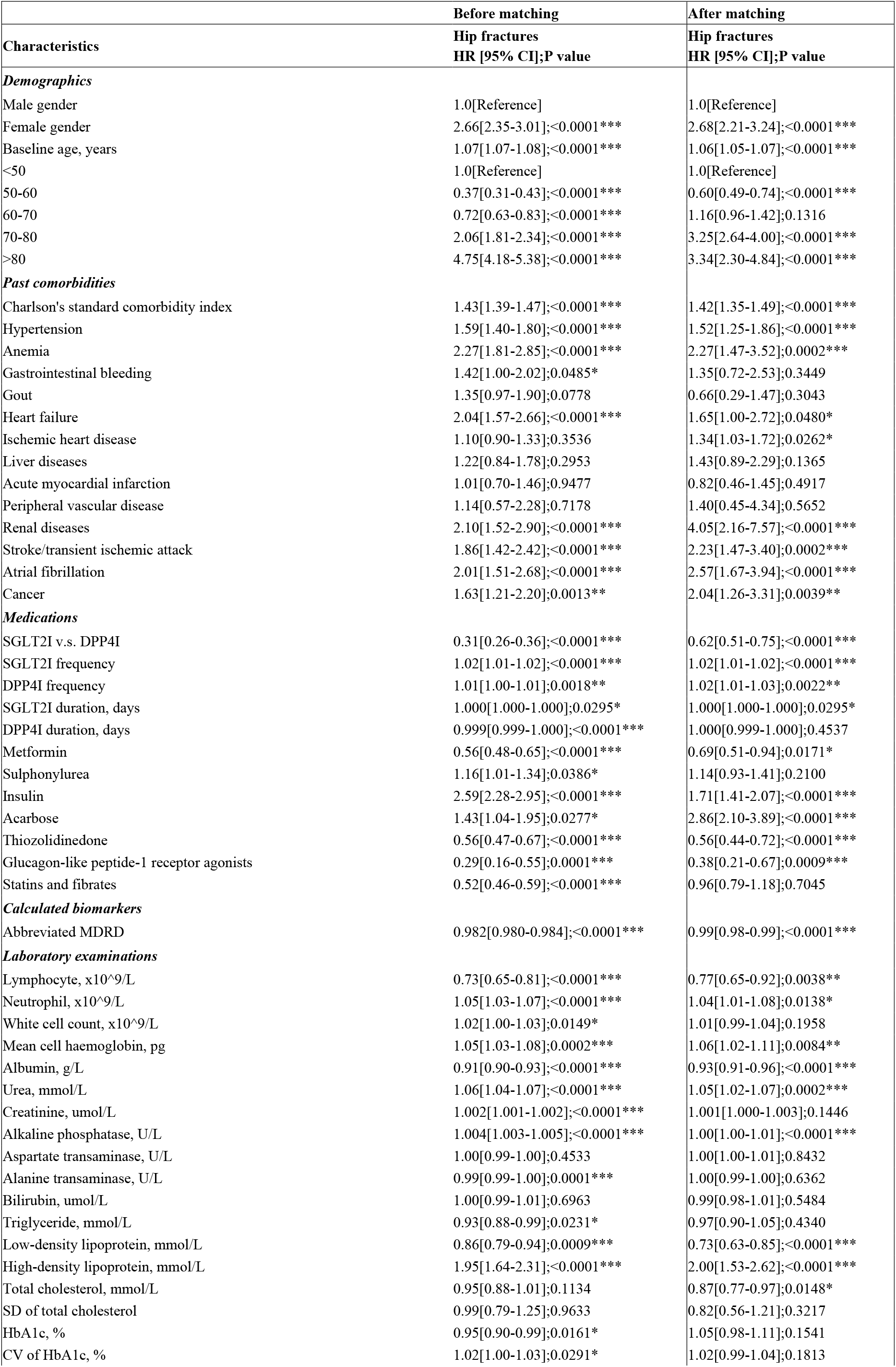
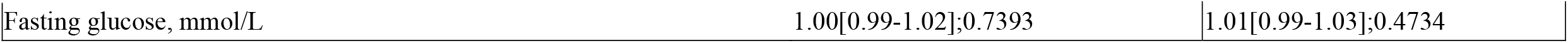
Univariable Cox analyses of significant risk factors for hip fractures before and after propensity score matching (1:1). * for p≤ 0.05, ** for p ≤ 0.01, *** for p ≤ 0.001; HR: hazard ratio; CI: confidence interval; SD: standard deviation; SGLT2I: sodium glucose cotransporter-2 inhibitor; DPP4I: dipeptidyl peptidase-4 inhibitor; CV: coefficient of variation.

**Table 3.**
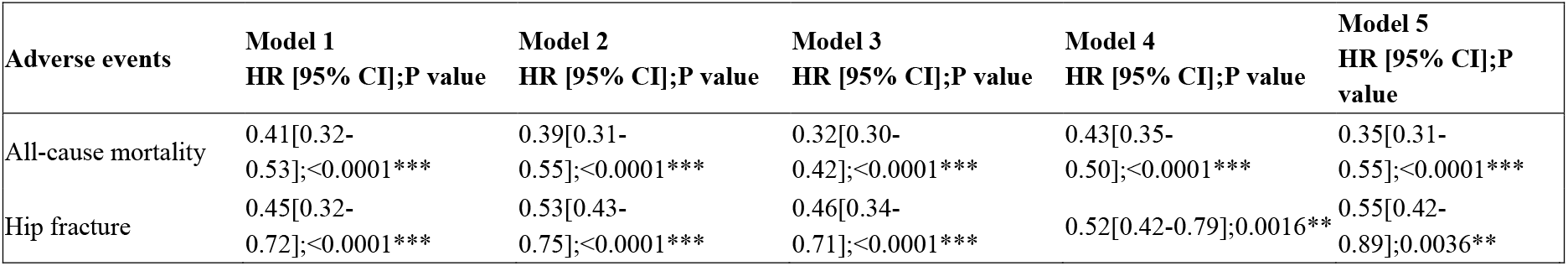
Multivariable Cox analysis for new-onset hip fractures and all-cause mortality in the matched cohort. * for p≤ 0.05, ** for p ≤ 0.01, *** for p ≤ 0.001; HR: hazard ratio; CI: confidence interval; SGLT2I: sodium glucose cotransporter-2 inhibitor; DPP4I: dipeptidyl peptidase-4 inhibitor. IR: incidence rate. Model 1 adjusted for significant demographics. Model 2 adjusted for significant demographics, and past comorbidities. Model 3 adjusted for significant demographics, past comorbidities, and non-SGLT2I/DPP4I medications. Model 4 adjusted for significant demographics, past comorbidities, non-SGLT2I/DPP4I medications, and laboratory results Model 5 adjusted for significant demographics, past comorbidities, non-SGLT2I/DPP4I medications, laboratory results, HbA1c and fasting glucose tests.

**Fig 2.**
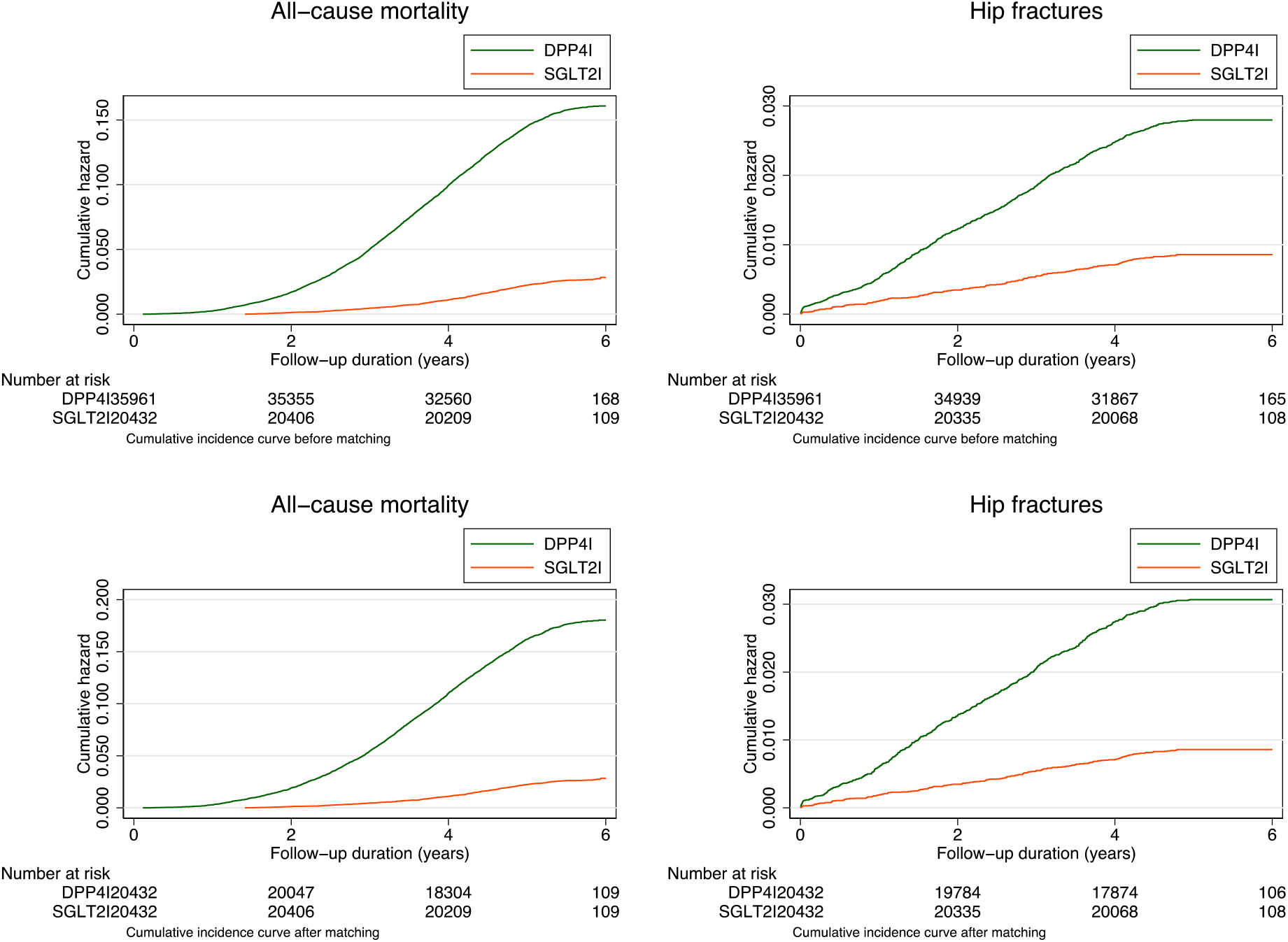
Cumulative incidence curves for new-onset hip fractures and all-cause mortality stratified by SGLT2I use v.s. DPP4I use before and after propensity score matching (1:1).

### Sensitivity analyses

Sensitivity analyses were performed to confirm the predictiveness of the models. The association between SGLT2I and hip fracture remains after a one year lag time was applied (HR: 0.69; 95% CI: 0.41-0.93; P=0.0020) **(Supplementary Table 4)**. Besides, SGLT2I was associated with lower risk of new-onset hip fractures in the cause-specific hazard (HR: 0.66; 95% CI: 0.45-0.90; P=0.0012) and the subdistribution hazard models (HR: 0.58; 95% CI: 0.52-0.89; P=0.0145) **(Supplementary Table 5)**. Lastly, using different propensity score approaches, SGLT2I was associated with lower risks of new-onset hip fracture in the propensity score stratification (HR: 0.62; 95% CI: 0.51-0.88; P=0.0002), inverse probability of treatment weighting (HR: 0.68, 95% CI: 0.42-0.97; P=0.0051), and stable inverse probability of treatment weighting model (HR: 0.72; 95% CI: 0.38-0.93; P=0.0011) **(Supplementary Table 6)**.

## Discussion

In this territory-wide retrospective cohort study, we used real-world data from routine clinical practice to compare the effects of SGLT2I and DPP4I on hip fracture. Our findings demonstrated that SGLT2I was associated with lower risks of new-onset hip fractures compared to DPP4I users, after propensity score matching. To the best of our knowledge, the present study is the first to show the fracture protective effect of SGLT2I, when compared to DPP4I directly.

### Potential underlying mechanisms

T2DM is known to be associated with an increased risk of fractures. It was previously suggested that the accumulation of AGEs would contribute to lower bone quality.^24^ Secondly, sclerostin expressed in osteocytes may also be responsible for the lower bone quality among T2DM patients.^25^ Last but not least, it was also suggested that the vitamin D level was lower among T2DM patients.^26^ However, the relationship between SGLT2I and fracture remains ambiguous. SGLT2I may result in hypercalciuria due to osmotic diuresis and reduce paracellular calcium reabsorption, thus, resulting in loss of calcium.^27^ It was also reported that SGLT2I may increase parathyroid hormone level, increase phosphate reabsorption level and decrease vitamin D activity.^28^ Furthermore, SGLT2I is associated with an increased level of bone reabsorption marker due to off-target effects on the bone.^29^ However, those findings may not necessarily translate to increased hip fracture risks in the real world, as previous studies did not find that SGLT2I increases fracture risks.^30, 31^

Meanwhile, the effects of DPP4I on bone fracture are also unclear. As GLP-1 is known to be beneficial to bone health, there were initial concerns regarding whether DPP4I would increase bone fracture risks. However, a meta-analysis in 2011 showed that DPP4I is associated with a reduced risk of fractures compared with placebo or other anti-diabetic medications^32^ while a more recent meta-analysis in 2019 showed DPP4I does not affect fracture risk.^33^ Our findings suggest that compared against DPP4I, SGLT2I is associated with a reduced risk of hip fractures. As the two anti-diabetic medications work via different mechanisms, more basic and comparative studies are needed to explain the reduced risk of hip fractures of SGLT2I compared to DPP4I.

### Comparison with previous studies

While SGLT2I has been shown to have better mortality outcome compared to DPP4I, the association of SGLT2I use with the risk of fractures remained as a subject of great debate. The Canagliflozin Cardiovascular Assessment Study (CANVAS) was the first trial to demonstrate an elevated risk of fracture in canagliflozin users compared to placebo.^34^ This prompted the United States Food and Drug Administration in 2015 to issue a warning about the increased risk of bone fractures among canagliflozin users.^11^ However, later analyses could not demonstrate any definitive explanation and suspected it to be a chance finding rather than an observed significant association.^35^ It should be noted that subsequent trials, ^36, 37^ observational studies ^38^ and meta-analyses ^30, 31^ showed inconsistent findings, with the majority establishing no significant association of SGLT2I use with an elevated risk of fracture ^39^.

To the best of our knowledge, the present study was the first to demonstrate a protective effect of SGLT2I on hip fractures under a direct comparison with DPP4I. Existing meta-analyses demonstrated that DPP4I has little, or even protective effects against the risk of fractures.^32, 40^ A previous study by Abrahami *et al*. ^41^ showed this protective effect only among canagliflozin users after the stratified analysis, although this was based on a small number of events (n = 10), in contrast to the present study where the findings were based on over 400 reported cases of new-onset hip fracture. Another study performed by Zhuo *et al*. found no association of SGLT2I with an increased risk of fracture in older adults with T2DM when compared with initiating a DPP4I or GLP-1 receptor agonist.^39^ However, the authors were unable to adjust for severity of diabetes and glycaemic control due to the unavailability of data on HbA1c, which was adjusted for in the present study. Some RCT of ganagliflozin also demonstrated that SGLT2I was not associated with increased risks of fractures.

### Clinical implications and the future

Whereas no conclusions have been reached so far regarding the relationship between SGLT2I and fracture risk, our study provides novel data to show that SGLT2I has favourable profiles on bone health and bone-related adverse outcomes when compared to DPP4I. Our findings hold great clinical relevance because osteoporotic fractures are a major reason for morbidity and mortality especially in the older population,^42^ and T2DM has been identified as an important risk factor for fractures.^43^ The current findings suggest that SGLT2I could serve as an antidiabetic treatment of choice in light of its effects on hip fracture. Our findings also highlight the safety profile of SGLT2I in terms of fracture risk, providing further reassurance that SGLT2I may not be associated with an increased risk of fractures as previously believed. However, further studies are needed to further elucidate the underlying mechanisms.

### Limitations

Several limitations should be noted for the present study. Firstly, given its observational nature, there is inherent information bias due to under-coding, coding errors, and missing data. Secondly, medication adherence can only be assessed indirectly through prescription refills, which are ultimately not a direct measurement of drug exposure. Thirdly, residual and post-baseline confounding may be present despite robust propensity-matching, particularly with the unavailability of information on lifestyle cardiovascular risk factors such as smoking, alcohol usage, BMI. Fourthly, the duration of drug exposure has not been controlled for, which may affect their risk against the study outcomes. Fifthly, the occurrence of new-onset hip fracture that did not require hospital admission was not accounted for. Lastly, our study’s retrospective design necessitates presentation of associations but not causal links between SGLT2I/DPP4I use and the risk of new-onset hip fractures.

## Conclusions

In this real-world cohort study, SGLT2I was associated with a lower risk of new-onset hip fractures compared to DPP4I after propensity score matching and adjustments. Further studies are needed to investigate the mechanism behind the observations and elucidate the causation.

## Data Availability

All data produced in the present study are available upon reasonable request to the authors

## Conflicts of Interest

None.

## Funding source

The author(s) received no funding for the research, authorship, and/or publication of this article.

## Availability of data and materials

An anonymised version without identifiable or personal information is available from the corresponding authors upon reasonable request for research purposes.

## Acknowledgements

None.

## Notes

declare that they have no conflict of interest.

### Competing Interest Statement

The authors have declared no competing interest.

### Funding Statement

This study did not receive any funding

### Author Declarations

This study was approved by The Joint Chinese University of Hong Kong/New Territories East Cluster Clinical Research Ethics Committee.

## References

1. Khan MAB, Hashim MJ, King JK, Govender RD, Mustafa H, Al Kaabi J. Epidemiology of Type 2 Diabetes - Global Burden of Disease and Forecasted Trends. J Epidemiol Glob Health. Mar 2020;10(1):107–111. doi:10.2991/jegh.k.191028.001

2. Viigimaa M, Sachinidis A, Toumpourleka M, Koutsampasopoulos K, Alliksoo S, Titma T. Macrovascular Complications of Type 2 Diabetes Mellitus. Curr Vasc Pharmacol. 2020;18(2):110–116. doi:10.2174/1570161117666190405165151

3. Fan Y, Wei F, Lang Y, Liu Y. Diabetes mellitus and risk of hip fractures: a meta-analysis. Osteoporos Int. Jan 2016;27(1):219–28. doi:10.1007/s00198-015-3279-7

4. Forsen L, Meyer HE, Midthjell K, Edna TH. Diabetes mellitus and the incidence of hip fracture: results from the Nord-Trondelag Health Survey. Diabetologia. Aug 1999;42(8):920–5. doi:10.1007/s001250051248

5. Tian W, Wu J, Tong T, et al. Diabetes and Risk of Post-Fragility Hip Fracture Outcomes in Elderly Patients. International Journal of Endocrinology. 2020/04/14 2020;2020:8146196. doi:10.1155/2020/8146196

6. Yang Y, Zhao C, Liang J, Yu M, Qu X. Effect of Dipeptidyl Peptidase-4 Inhibitors on Bone Metabolism and the Possible Underlying Mechanisms. Mini Review. Frontiers in Pharmacology. 2017-July-25 2017;8doi:10.3389/fphar.2017.00487

7. Hou WH, Chang KC, Li CY, Ou HT. Dipeptidyl peptidase-4 inhibitor use is associated with decreased risk of fracture in patients with type 2 diabetes: a population-based cohort study. Br J Clin Pharmacol. Sep 2018;84(9):2029–2039. doi:10.1111/bcp.13636

8. Gamble J-M, Donnan JR, Chibrikov E, Twells LK, Midodzi WK, Majumdar SR. The risk of fragility fractures in new users of dipeptidyl peptidase-4 inhibitors compared to sulfonylureas and other anti-diabetic drugs: A cohort study. Diabetes Research and Clinical Practice. 2018/02/01/ 2018;136:159–167. doi:https://doi.org/10.1016/j.diabres.2017.12.008

9. Montagnani A, Gonnelli S. Antidiabetic therapy effects on bone metabolism and fracture risk. Diabetes Obes Metab. Sep 2013;15(9):784–91. doi:10.1111/dom.12077

10. Sanz C, Vázquez P, Blázquez C, Barrio PA, Alvarez Mdel M, Blázquez E. Signaling and biological effects of glucagon-like peptide 1 on the differentiation of mesenchymal stem cells from human bone marrow. Am J Physiol Endocrinol Metab. Mar 2010;298(3):E634–43. doi:10.1152/ajpendo.00460.2009

11. FDA Drug Safety Communication: FDA revises label of diabetes drug canagliflozin (Invokana, Invokamet) to include updates on bone fracture risk and new information on decreased bone mineral density. The U.S. Food and Drug Administration (FDA). 2021. https://www.fda.gov/drugs/drug-safety-and-availability/fda-drug-safety-communication-fda-revises-label-diabetes-drug-canagliflozin-invokana-invokamet

12. Lou Y, Yu Y, Duan J, et al. Sodium-glucose cotransporter 2 inhibitors and fracture risk in patients with type 2 diabetes mellitus: a meta-analysis of randomized controlled trials. Ther Adv Chronic Dis. 2020;11:2040622320961599. doi:10.1177/2040622320961599

13. Donnan JR, Grandy CA, Chibrikov E, et al. Comparative safety of the sodium glucose co-transporter 2 (SGLT2) inhibitors: a systematic review and meta-analysis. BMJ Open. Feb 1 2019;9(1):e022577. doi:10.1136/bmjopen-2018-022577

14. Zhuo M, Hawley CE, Paik JM, et al. Association of Sodium-Glucose Cotransporter–2 Inhibitors With Fracture Risk in Older Adults With Type 2 Diabetes. JAMA Network Open. 2021;4(10):e2130762–e2130762. doi:10.1001/jamanetworkopen.2021.30762

15. Watts NB, Bilezikian JP, Usiskin K, et al. Effects of Canagliflozin on Fracture Risk in Patients With Type 2 Diabetes Mellitus. J Clin Endocrinol Metab. Jan 2016;101(1):157–66. doi:10.1210/jc.2015-3167

16. Mui JV, Zhou J, Lee S, et al. Sodium-Glucose Cotransporter 2 (SGLT2) Inhibitors vs. Dipeptidyl Peptidase-4 (DPP4) Inhibitors for New-Onset Dementia: A Propensity Score-Matched Population-Based Study With Competing Risk Analysis. Front Cardiovasc Med. 2021;8:747620. doi:10.3389/fcvm.2021.747620

17. Zhou J, Lee S, Leung KSK, et al. Incident heart failure and myocardial infarction in sodium-glucose cotransporter-2 vs. dipeptidyl peptidase-4 inhibitor users. ESC Heart Fail. Feb 7 2022;doi:10.1002/ehf2.13830

18. Zhou J, Lee S, Liu X, et al. Hip fractures risks in edoxaban versus warfarin users: A propensity score-matched population-based cohort study with competing risk analyses. Bone. 2022/03/01/ 2022;156:116303. doi:https://doi.org/10.1016/j.bone.2021.116303

19. Zhou J, Li H, Chang C, et al. The association between blood pressure variability and hip or vertebral fracture risk: A population-based study. Bone. Sep 2021;150:116015. doi:10.1016/j.bone.2021.116015

20. Soliman AR, Fathy A, Khashab S, Shaheen N. Comparison of abbreviated modification of diet in renal disease formula (aMDRD) and the Cockroft-Gault adjusted for body surface (aCG) equations in stable renal transplant patients and living kidney donors. Ren Fail. 2013;35(1):94–7. doi:10.3109/0886022x.2012.731970

21. Austin PC. An Introduction to Propensity Score Methods for Reducing the Effects of Confounding in Observational Studies. Multivariate Behav Res. 2011;46(3):399–424. doi:10.1080/00273171.2011.568786

22. Austin PC, Stuart EA. Moving towards best practice when using inverse probability of treatment weighting (IPTW) using the propensity score to estimate causal treatment effects in observational studies. Stat Med. Dec 10 2015;34(28):3661–79. doi:10.1002/sim.6607

23. Avagyan V, Vansteelandt S. Stable inverse probability weighting estimation for longitudinal studies. Scandinavian Journal of Statistics. 2021;48(3):1046–1067. doi:https://doi.org/10.1111/sjos.12542

24. Saito M, Fujii K, Soshi S, Tanaka T. Reductions in degree of mineralization and enzymatic collagen cross-links and increases in glycation-induced pentosidine in the femoral neck cortex in cases of femoral neck fracture. Osteoporosis International. 2006/07/01 2006;17(7):986–995. doi:10.1007/s00198-006-0087-0

25. Wang N, Xue P, Wu X, Ma J, Wang Y, Li Y. Role of sclerostin and dkk1 in bone remodeling in type 2 diabetic patients. Endocrine Research. 2018/01/02 2018;43(1):29–38. doi:10.1080/07435800.2017.1373662

26. Kostoglou-Athanassiou I, Athanassiou P, Gkountouvas A, Kaldrymides P. Vitamin D and glycemic control in diabetes mellitus type 2. Ther Adv Endocrinol Metab. Aug 2013;4(4):122–8. doi:10.1177/2042018813501189

27. Edwards A, Bonny O. A model of calcium transport and regulation in the proximal tubule. Am J Physiol Renal Physiol. Oct 1 2018;315(4):F942–f953. doi:10.1152/ajprenal.00129.2018

28. Vinke JSJ, Heerspink HJL, de Borst MH. Effects of sodium glucose cotransporter 2 inhibitors on mineral metabolism in type 2 diabetes mellitus. Curr Opin Nephrol Hypertens. Jul 2019;28(4):321–327. doi:10.1097/MNH.0000000000000505

29. Bilezikian JP, Watts NB, Usiskin K, et al. Evaluation of Bone Mineral Density and Bone Biomarkers in Patients With Type 2 Diabetes Treated With Canagliflozin. J Clin Endocrinol Metab. Jan 2016;101(1):44–51. doi:10.1210/jc.2015-1860

30. Ruanpeng D, Ungprasert P, Sangtian J, Harindhanavudhi T. Sodium-glucose cotransporter 2 (SGLT2) inhibitors and fracture risk in patients with type 2 diabetes mellitus: A meta-analysis. Diabetes Metab Res Rev. Sep 2017;33(6)doi:10.1002/dmrr.2903

31. Azharuddin M, Adil M, Ghosh P, Sharma M. Sodium-glucose cotransporter 2 inhibitors and fracture risk in patients with type 2 diabetes mellitus: A systematic literature review and Bayesian network meta-analysis of randomized controlled trials. Diabetes Res Clin Pract. Dec 2018;146:180–190. doi:10.1016/j.diabres.2018.10.019

32. Monami M, Dicembrini I, Antenore A, Mannucci E. Dipeptidyl peptidase-4 inhibitors and bone fractures: a meta-analysis of randomized clinical trials. Diabetes Care. Nov 2011;34(11):2474–6. doi:10.2337/dc11-1099

33. Chen Q, Liu T, Zhou H, Peng H, Yan C. Risk of Fractures Associated with Dipeptidyl Peptidase-4 Inhibitor Treatment: A Systematic Review and Meta-Analysis of Randomized Controlled Trials. Diabetes Therapy. 2019/10/01 2019;10(5):1879–1892. doi:10.1007/s13300-019-0668-5

34. Neal B, Perkovic V, Mahaffey KW, et al. Canagliflozin and Cardiovascular and Renal Events in Type 2 Diabetes. N Engl J Med. Aug 17 2017;377(7):644–657. doi:10.1056/NEJMoa1611925

35. Zhou Z, Jardine M, Perkovic V, et al. Canagliflozin and fracture risk in individuals with type 2 diabetes: results from the CANVAS Program. Diabetologia. Oct 2019;62(10):1854–1867. doi:10.1007/s00125-019-4955-5

36. Zinman B, Wanner C, Lachin JM, et al. Empagliflozin, Cardiovascular Outcomes, and Mortality in Type 2 Diabetes. N Engl J Med. Nov 26 2015;373(22):2117–28. doi:10.1056/NEJMoa1504720

37. Heerspink HJL, Stefansson BV, Correa-Rotter R, et al. Dapagliflozin in Patients with Chronic Kidney Disease. N Engl J Med. Oct 8 2020;383(15):1436–1446. doi:10.1056/NEJMoa2024816

38. Hidayat K, Du X, Shi BM. Risk of fracture with dipeptidyl peptidase-4 inhibitors, glucagon-like peptide-1 receptor agonists, or sodium-glucose cotransporter-2 inhibitors in real-world use: systematic review and meta-analysis of observational studies. Osteoporos Int. Oct 2019;30(10):1923–1940. doi:10.1007/s00198-019-04968-x

39. Zhuo M, Hawley CE, Paik JM, et al. Association of Sodium-Glucose Cotransporter-2 Inhibitors With Fracture Risk in Older Adults With Type 2 Diabetes. JAMA Netw Open. Oct 1 2021;4(10):e2130762. doi:10.1001/jamanetworkopen.2021.30762

40. Fu J, Zhu J, Hao Y, Guo C, Zhou Z. Dipeptidyl peptidase-4 inhibitors and fracture risk: an updated meta-analysis of randomized clinical trials. Sci Rep. Jul 7 2016;6:29104. doi:10.1038/srep29104

41. Abrahami D, Douros A, Yin H, Yu OHY, Azoulay L. Sodium-Glucose Cotransporter 2 Inhibitors and the Risk of Fractures Among Patients With Type 2 Diabetes. Diabetes Care. Sep 2019;42(9):e150–e152. doi:10.2337/dc19-0849

42. Johnell O, Kanis JA. An estimate of the worldwide prevalence and disability associated with osteoporotic fractures. Osteoporos Int. Dec 2006;17(12):1726–33. doi:10.1007/s00198-006-0172-4

43. Moseley KF. Type 2 diabetes and bone fractures. Curr Opin Endocrinol Diabetes Obes. Apr 2012;19(2):128–35. doi:10.1097/MED.0b013e328350a6e1

