## Supplementary Appendix for "Lower risks of sodium glucose cotransporter 2 (SGLT2) inhibitors compared to dipeptidyl peptidase-4 (DPP4) inhibitors for new-onset hip fracture risks in patients with type-2 diabetes: A propensity score-matched study with competing risk analysis"

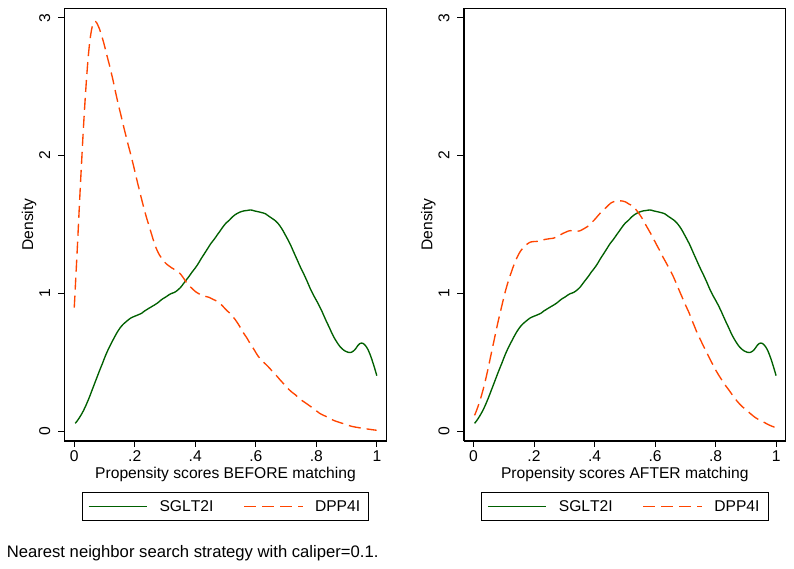

**Supplementary Figure 1. Propensity score matching comparisons and proportional hazard assumption checking with parallel lines for SGLT2I v.s. DPP4I before and after 1:1 matching with nearest neighbor search strategy using a caliper of 0..1**

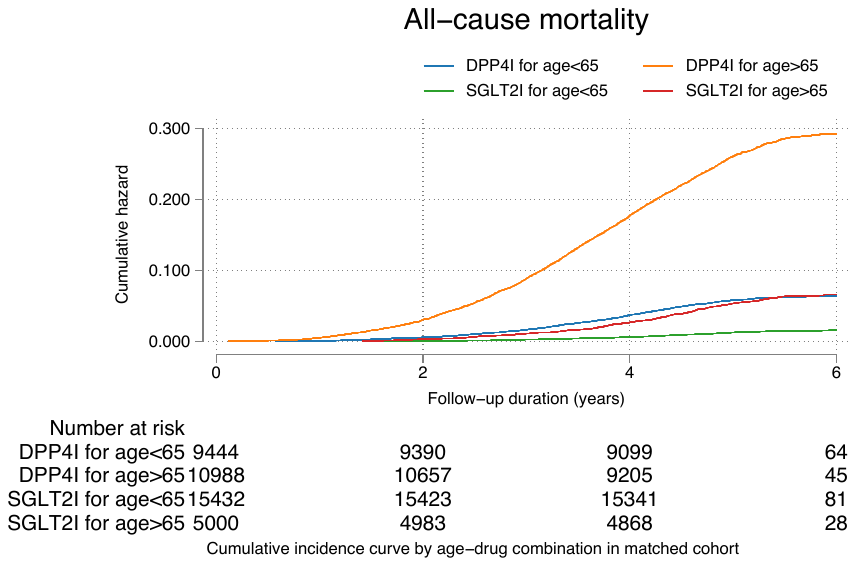

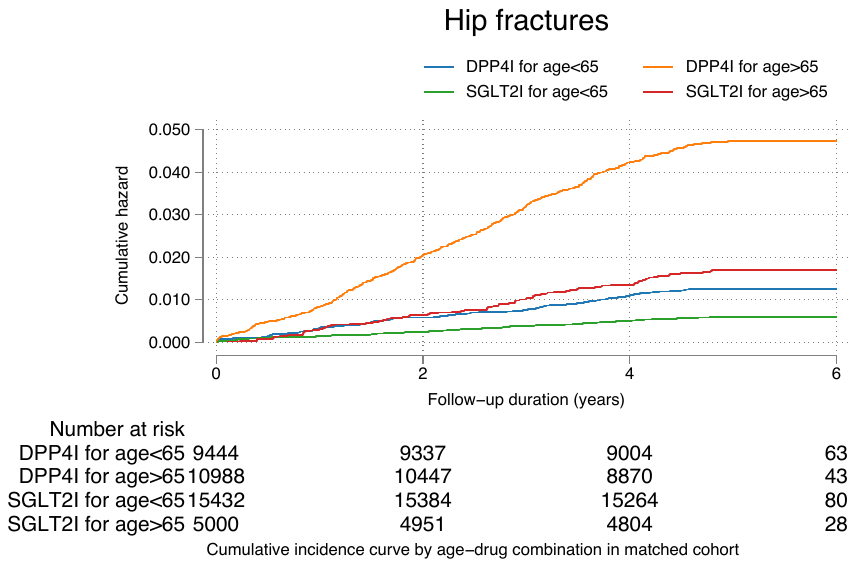

**Supplementary Figure 2. Subgroup analysis. Cumulative incidence curves for new onset hip fractures and all-cause mortality stratified by combinations of age and SGLT2I use v.s. DPP4I use in the matched cohort (1:1).**

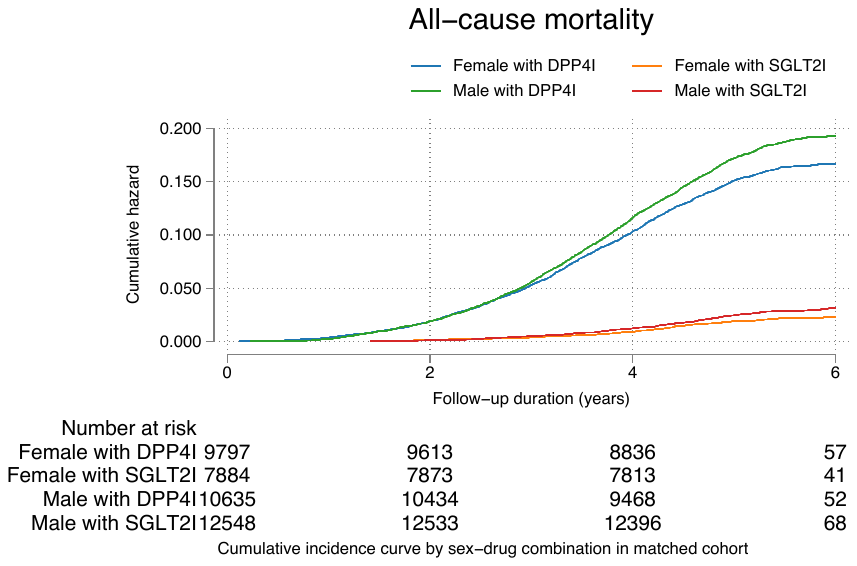

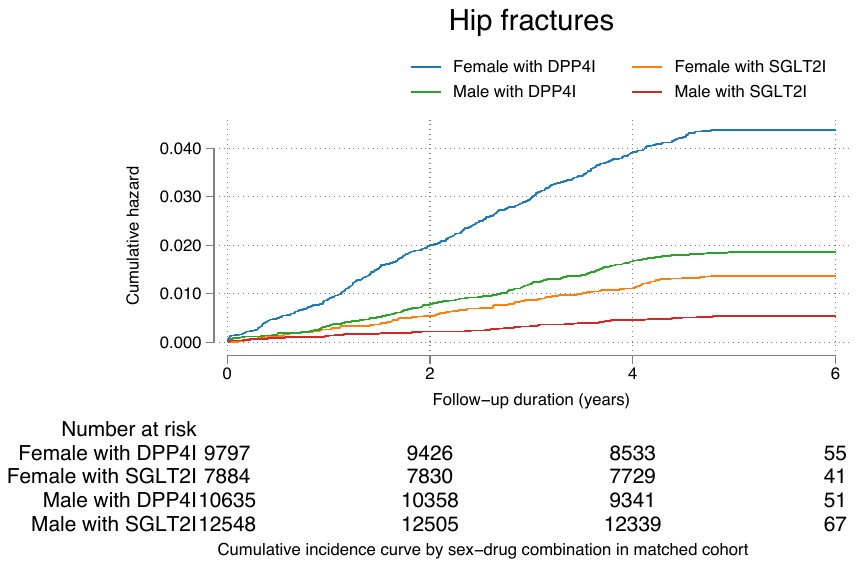

**Supplementary Figure 3. Subgroup analysis. Cumulative incidence curves for new onset hip fractures and all-cause mortality stratified by combinations of gender and SGLT2I use v.s. DPP4I use in the matched cohort (1:1).**

**Supplementary Table 1. ICD-9 codes for comorbidities and ICD10 codes for outcomes.**

Diabetes mellitus 250	250.01	250.02	250.03	250.1	250.11	250.12	250.13	250.2	250.21	250.22	250.23	250.3	250.31	250.32	250.33	250.4	250.41	250.42	250.43	250.5	250.51	250.52	250.53	250.6	250.61	250.62	250.63	250.7	250.71	250.72	250.73	250.8	250.81	250.82	250.83	250.9	250.91	250.92	250.93

Renal diseases	582	582	582.1	582.2	582.4	582.8	582.81	582.89	582.9	583	583	583.1	583.2	583.4	583.6	583.7	585	585.1	585.2	585.3	585.4	585.5	585.6	585.9	586	588	588	588.1	588.8	588.81	588.89	588.9

Acute myocardial infarction 410	410.01	410.02	410.1	410.11	410.12	410.2	410.21	410.22	410.3	410.31	410.32	410.4	410.41	410.42	410.5	410.51	410.52	410.6	410.61	410.62	410.7	410.71	410.72	410.8	410.81	410.82	410.9	410.91	410.92

Hypertension	401	401.1	401.9	402	402.01	402.1	402.11	402.9	402.91	403	403.01	403.1	403.11	403.9	403.91	404	404.01	404.02	404.03	404.1	404.11	404.12	404.13	404.9	404.91	404.92	404.93	405	405.01	405.09	405.1	405.11	405.19	405.9	405.91	405.99	437.2

Heart failure	428	428	428.1	428.2	428.2	428.21	428.22	428.23	428.3	428.3	428.31	428.32	428.33	428.4	428.4	428.41	428.42	428.43	428.9	398.91	402.01	402.11	402.91	404.01	404.03	404.11	404.13	404.91	404.93

Atrial fibrillation	427.31	429.4

Liver diseases	456	456.1	456.2	572.2	572.3	572.4	572.8	571.4	571.5	571.6

Chronic obstructive pulmonary disease	490	491	492	493	494	495	496	491.1	491.2	491.21	491.22	491.8	491.9	492.8	493.01	493.02	493.1	493.11	493.12	493.2	493.21	493.22	493.8	493.81	493.82	493.9	493.91	493.92	494.1	495.1	495.2	495.3	495.4	495.5	495.6	495.7	495.8	495.9

Peripheral vascular disease	250.7	443.9	443	443.1	443.2	443.21	443.22	443.23	443.24	443.29	443.8	443.81	443.82	443.89	441	443.9	785.4	V43.4

Stroke/transient ischemic attack	435	435.1	435.2	435.3	435.8	435.9	433.81	433.91	434	436	437	437.1	433.31	433.01	434.01	434.1	434.11	434.9	434.91	437.2	437.3	437.4	437.5	437.6	437.7	437.8	437.9	430	431	432	432.1	432.9

Ischemic heart disease	410.01	410.02	410.1	410.11	410.12	410.2	410.21	410.22	410.3	410.31	410.32	410.4	410.41	410.42	410.5	410.51	410.52	410.6	410.61	410.62	410.7	410.71	410.72	410.8	410.81	410.82	410.9	410.91	410.92	411	411.1	411.8	411.81	411.89	413	413.1	413.9	414	414.01	414.02	414.03	414.04	414.05	414.06	414.07	414.1	414.11	414.12	414.19	414.2	414.3	414.4	414.8	414.9	410	412

Cancer 140	140.1	140.3	140.4	140.5	140.6	140.8	140.9	141	141.1	141.2	141.3	141.4	141.5	141.6	141.8	141.9	142	142.1	142.2	142.8	142.9	143	143.1	143.8	143.9	144	144.1	144.8	144.9	145	145.1	145.2	145.3	145.4	145.5	145.6	145.8	145.9	146	146.1	146.2	146.3	146.4	146.5	146.6	146.7	146.8	146.9	147	147.1	147.2	147.3	147.8	147.9	148	148.1	148.2	148.3	148.8	148.9	149	149.1	149.8	149.9	150	150.1	150.2	150.3	150.4	150.5	150.8	150.9	151	151.1	151.2	151.3	151.4	151.5	151.6	151.8	151.9	152	152.1	152.2	152.3	152.8	152.9	153	153.1	153.2	153.3	153.4	153.5	153.6	153.7	153.8	153.9	154	154.1	154.2	154.3	154.8	155	155.1	155.2	156	156.1	156.2	156.8	156.9	157	157.1	157.2	157.3	157.4	157.8	157.9	158	158.8	158.9	159	159.1	159.8	159.9	160	160.1	160.2	160.3	160.4	160.5	160.8	160.9	161	161.1	161.2	161.3	161.8	161.9	162	162.2	162.3	162.4	162.5	162.8	162.9	163	163.1	163.8	163.9	164	164.1	164.2	164.3	164.8	164.9	165	165.8	165.9	170	170.1	170.2	170.3	170.4	170.5	170.6	170.7	170.8	170.9	171	171.2	171.3	171.4	171.5	171.6	171.7	171.8	171.9	172	172.1	172.2	172.3	172.4	172.5	172.6	172.7	172.8	172.9 173	173.01	173.02	173.09	173.1	173.11	173.12	173.19	173.2	173.21	173.22	173.29	173.3	173.31	173.32	173.39	173.4	173.41	173.42	173.49	173.5	173.51	173.52	173.59	173.6	173.61	173.62	173.69	173.7	173.71	173.72	173.79	173.8	173.81	173.82	173.89	173.9	173.91	173.92	173.99	174	174.1	174.2	174.3	174.4	174.5	174.6	174.8	174.9	175	175.9	176	176.1	176.2	176.3	176.4	176.5	176.8	176.9	179	180	180.1	180.8	180.9	181	182	182.1	182.8	183	183.2	183.3	183.4	183.5	183.8	183.9	184	184.1	184.2	184.3	184.4	184.8	184.9	185	186	186.9	187	187.1	187.2	187.3	187.4	187.5	187.6	187.7	187.8	187.9	188	188.1	188.2	188.3	188.4	188.5	188.6	188.7	188.8	188.9	189	189.1	189.2	189.3	189.4	189.8	189.9	190	190.1	190.2	190.3	190.4	190.5	190.6	190.7	190.8	190.9	191	191.1	191.2	191.3	191.4	191.5	191.6	191.7	191.8	191.9	192	192.1	192.2	192.3	192.8	192.9	193	194	194.1	194.3	194.4	194.5	194.6	194.8	194.9	195	195.1	195.2	195.3	195.4	195.5	195.8	200	200.01	200.02	200.03	200.04	200.05	200.06	200.07	200.08	200.1	200.11	200.12	200.13	200.14	200.15	200.16	200.17	200.18	200.2	200.21	200.22	200.23	200.24	200.25	200.26	200.27	200.28	200.3	200.31	200.32	200.33	200.34	200.35	200.36	200.37	200.38	200.4	200.41	200.42	200.43	200.44	200.45	200.46	200.47	200.48	200.5	200.51	200.52	200.53	200.54	200.55	200.56	200.57	200.58	200.6	200.61	200.62	200.63	200.64	200.65	200.66	200.67	200.68	200.7	200.71	200.72	200.73	200.74	200.75	200.76	200.77	200.78	200.8	200.81	200.82	200.83	200.84	200.85	200.86	200.87	200.88	201	201.01	201.02	201.03	201.04	201.05	201.06	201.07	201.08	201.1	201.11	201.12	201.13	201.14	201.15	201.16	201.17	201.18	201.2	201.21	201.22	201.23	201.24	201.25	201.26	201.27	201.28	201.4	201.41	201.42	201.43	201.44	201.45	201.46	201.47	201.48	201.5	201.51	201.52	201.53	201.54	201.55	201.56	201.57	201.58	201.6	201.61	201.62	201.63	201.64	201.65	201.66	201.67	201.68	201.7	201.71	201.72	201.73	201.74	201.75	201.76	201.77	201.78	201.9	201.91	201.92	201.93	201.94	201.95	201.96	201.97	201.98	202	202.01	202.02	202.03	202.04	202.05	202.06	202.07	202.08	202.1	202.11	202.12	202.13	202.14	202.15	202.16	202.17	202.18	202.2	202.21	202.22	202.23	202.24	202.25	202.26	202.27	202.28	202.3	202.31	202.32	202.33	202.34	202.35	202.36	202.37	202.38	202.4	202.41	202.42	202.43	202.44	202.45	202.46	202.47	202.48	202.5	202.51	202.52	202.53	202.54	202.55	202.56	202.57	202.58	202.6	202.61	202.62	202.63	202.64	202.65	202.66	202.67	202.68	202.7	202.71	202.72	202.73	202.74	202.75	202.76	202.77	202.78	202.8	202.81	202.82	202.83	202.84	202.85	202.86	202.87	202.88	202.9	202.91	202.92	202.93	202.94	202.95	202.96	202.97	202.98	203	203.01	203.02	203.1	203.11	203.12	203.8	203.81	203.82	204	204.01	204.02	204.1	204.11	204.12	204.2	204.21	204.22	204.8	204.81	204.82	204.9	204.91	204.92	205	205.01	205.02	205.1	205.11	205.12	205.2	205.21	205.22	205.3	205.31	205.32	205.8	205.81	205.82	205.9	205.91	205.92	206	206.01	206.02	206.1	206.11	206.12	206.2	206.21	206.22	206.8	206.81	206.82	206.9	206.91	206.92	207	207.01	207.02	207.1	207.11	207.12	207.2	207.21	207.22	207.8	207.81	207.82	208	208.01	208.02	208.1	208.11	208.12	208.2	208.21	208.22	208.8	208.81	208.82	208.9	208.91	208.92 196	196.1	196.2	196.3	196.5	196.6	196.8	196.9	197	197.1	197.2	197.3	197.4	197.5	197.6	197.7	197.8	198	198.1	198.2	198.3	198.4	198.5	198.6	198.7	198.8	198.81	198.82	198.89	199	199.1

Hypertension	401	401.1	401.9	402	402.01	402.1	402.11	402.9	402.91	403	403.01	403.1	403.11	403.9	403.91	404	404.01	404.02	404.03	404.1	404.11	404.12	404.13	404.9	404.91	404.92	404.93	405	405.01	405.09	405.1	405.11	405.19	405.9	405.91	405.99	437.2

Anemia	280	280.1	280.8	280.9	281	281.1	281.2	281.3	281.4	281.8	281.9	282.2	282.3	282.8	282.9	283	283.1	283.11	283.19	283.2	283.9	284	284.01	284.09	284.1	284.11	284.12	284.19	284.81	284.9	285	285.1	285.2	285.21	285.22	285.29	285.3	285.8	285.9

Overweight	278	278	278	278.01	278.02	278.03	278.1	278.2	278.3	278.4	278.8

Gout	274	274.01	274.02	274.03	274.1	274.11	274.19	274.8	274.81	274.82	274.89	274.9

Cardiovascular mortality I00-I09, I11, I13, I20-I51

**Supplementary Table 2. Calculations for variability measures.**

| **Variability measure** | **Definition** |
| --- | --- |
| Standard deviation | $\sqrt{\frac{1}{Number of measurements}\sum_{i=1}^{Number of measurements} {({test}_{i}-individual mean)}^{2}}$ |
| Coefficient of variation | $\frac{SD}{individual mean}$ |

.

**Supplementary Table 3. Univariable Cox regression models to predict mortality before and after 1:1 matching.**

* for p≤ 0.05, ** for p ≤ 0.01, *** for p ≤ 0.001; HR: hazard ratio; CI: confidence interval; SD: standard deviation; SGLT2I: sodium glucose cotransporter-2 inhibitor; DPP4I: dipeptidyl peptidase-4 inhibitor; CV: coefficient of variation.

|  | **Before matching** | **After matching** |
| --- | --- | --- |
| **Characteristics** | **All-cause mortality**  **HR [95% CI];P value** | **All-cause mortality**  **HR [95% CI];P value** |
| ***Demographics*** |  |  |
| Male gender | 1.0[Reference] | 1.0[Reference] |
| Female gender | 0.96[0.91-1.01];0.1281 | 0.87[0.80-0.96];0.0037** |
| Baseline age, years | 1.09[1.09-1.10];<0.0001*** | 1.080[1.075-1.084];<0.0001*** |
| <50 | 1.0[Reference] | 1.0[Reference] |
| 50-60 | 0.27[0.25-0.30];<0.0001*** | 0.47[0.42-0.52];<0.0001*** |
| 60-70 | 0.58[0.55-0.62];<0.0001*** | 1.14[1.04-1.25];0.0070** |
| 70-80 | 2.09[1.97-2.21];<0.0001*** | 3.15[2.86-3.48];<0.0001*** |
| >80 | 6.73[6.38-7.10];<0.0001*** | 7.21[6.32-8.23];<0.0001*** |
| ***Past comorbidities*** |  |  |
| Charlson's standard comorbidity index | 1.54[1.52-1.55];<0.0001*** | 1.56[1.53-1.59];<0.0001*** |
| Hypertension | 1.87[1.77-1.97];<0.0001*** | 1.65[1.50-1.81];<0.0001*** |
| Anemia | 2.96[2.71-3.25];<0.0001*** | 2.35[1.91-2.88];<0.0001*** |
| Gastrointestinal bleeding | 1.87[1.63-2.15];<0.0001*** | 1.94[1.51-2.49];<0.0001*** |
| Gout | 2.46[2.20-2.76];<0.0001*** | 2.00[1.60-2.51];<0.0001*** |
| Heart failure | 4.08[3.73-4.46];<0.0001*** | 4.77[4.10-5.54];<0.0001*** |
| Ischemic heart disease | 1.59[1.48-1.72];<0.0001*** | 1.90[1.71-2.12];<0.0001*** |
| Liver diseases | 1.04[0.87-1.25];0.6350 | 0.97[0.75-1.27];0.8426 |
| Acute myocardial infarction | 2.01[1.79-2.27];<0.0001*** | 2.69[2.29-3.15];<0.0001*** |
| Peripheral vascular disease | 3.79[3.18-4.53];<0.0001*** | 5.99[4.56-7.87];<0.0001*** |
| Renal diseases | 4.42[3.98-4.92];<0.0001*** | 3.73[2.74-5.08];<0.0001*** |
| Stroke/transient ischemic attack | 2.26[2.03-2.53];<0.0001*** | 2.20[1.80-2.69];<0.0001*** |
| Atrial fibrillation | 3.06[2.75-3.40];<0.0001*** | 3.58[3.00-4.27];<0.0001*** |
| Overweight | 0.47[0.30-0.74];0.0010** | 0.48[0.29-0.80];0.0045** |
| Alcohol dependence | 4.61[3.42-6.22];<0.0001*** | 5.32[2.94-9.61];<0.0001*** |
| Cancer | 2.35[2.10-2.63];<0.0001*** | 3.22[2.66-3.88];<0.0001*** |
| ***Medications*** |  |  |
| SGLT2I v.s. DPP4I | 0.17[0.15-0.18];<0.0001*** | 0.34[0.31-0.38];<0.0001*** |
| SGLT2I frequency | 1.02[1.01-1.02];<0.0001*** | 1.02[1.01-1.02];<0.0001*** |
| DPP4I frequency | 0.99[0.99-1.00];0.0185* | 1.03[1.03-1.04];<0.0001*** |
| SGLT2I duration, days | 1.000[1.000-1.000];0.0057** | 1.000[1.000-1.000];0.0057** |
| DPP4I duration, days | 0.999[0.999-0.999];<0.0001*** | 0.998[0.998-0.998];<0.0001*** |
| Metformin | 0.27[0.26-0.29];<0.0001*** | 0.39[0.35-0.44];<0.0001*** |
| Sulphonylurea | 1.05[0.99-1.12];0.0903 | 1.08[0.98-1.20];0.1051 |
| Insulin | 5.13[4.79-5.49];<0.0001*** | 4.52[4.04-5.07];<0.0001*** |
| Acarbose | 1.07[0.91-1.25];0.4209 | 1.93[1.62-2.28];<0.0001*** |
| Thiozolidinedone | 0.37[0.33-0.40];<0.0001*** | 0.51[0.45-0.57];<0.0001*** |
| Glucagon-like peptide-1 receptor agonists | 0.13[0.09-0.19];<0.0001*** | 0.25[0.18-0.35];<0.0001*** |
| Statins and fibrates | 0.31[0.29-0.33];<0.0001*** | 0.77[0.70-0.85];<0.0001*** |
| ***Calculated biomarkers*** |  |  |
| Abbreviated MDRD | 0.970[0.969-0.971];<0.0001*** | 0.98[0.97-0.98];<0.0001*** |
| ***Laboratory examinations*** |  |  |
| Mean corpuscular volume, fL | 1.02[1.02-1.03];<0.0001*** | 1.03[1.02-1.04];<0.0001*** |
| Eosinophil, x10^9/L | 1.09[0.98-1.21];0.1197 | 0.62[0.43-0.88];0.0080** |
| Lymphocyte, x10^9/L | 0.51[0.48-0.53];<0.0001*** | 0.65[0.60-0.71];<0.0001*** |
| Neutrophil, x10^9/L | 1.05[1.04-1.06];<0.0001*** | 1.06[1.05-1.07];<0.0001*** |
| White cell count, x10^9/L | 1.02[1.01-1.02];<0.0001*** | 1.02[1.01-1.03];<0.0001*** |
| Mean cell haemoglobin, pg | 1.06[1.05-1.07];<0.0001*** | 1.07[1.04-1.09];<0.0001*** |
| Platelet, x10^9/L | 0.997[0.996-0.997];<0.0001*** | 0.997[0.996-0.997];<0.0001*** |
| Red cell count, x10^12/L | 0.39[0.38-0.41];<0.0001*** | 0.44[0.40-0.48];<0.0001*** |
| Potassium, mmol/L | 1.14[1.08-1.21];<0.0001*** | 0.98[0.88-1.09];0.7087 |
| Albumin, g/L | 0.86[0.86-0.87];<0.0001*** | 0.86[0.85-0.87];<0.0001*** |
| Sodium, mmol/L | 0.95[0.94-0.96];<0.0001*** | 0.92[0.91-0.94];<0.0001*** |
| Urea, mmol/L | 1.089[1.086-1.092];<0.0001*** | 1.08[1.07-1.09];<0.0001*** |
| Protein, g/L | 0.95[0.94-0.95];<0.0001*** | 0.97[0.96-0.98];<0.0001*** |
| Creatinine, umol/L | 1.003[1.002-1.003];<0.0001*** | 1.003[1.003-1.003];<0.0001*** |
| Alkaline phosphatase, U/L | 1.00[1.00-1.01];<0.0001*** | 1.006[1.005-1.006];<0.0001*** |
| Aspartate transaminase, U/L | 1.000[1.000-1.001];0.3308 | 1.000[0.997-1.002];0.8460 |
| Alanine transaminase, U/L | 0.99[0.98-0.99];<0.0001*** | 0.99[0.99-1.00];<0.0001*** |
| Bilirubin, umol/L | 1.00[0.99-1.00];0.0907 | 1.01[1.00-1.02];0.0013** |
| Triglyceride, mmol/L | 0.94[0.92-0.96];<0.0001*** | 0.88[0.84-0.93];<0.0001*** |
| Low-density lipoprotein, mmol/L | 0.93[0.90-0.97];0.0004*** | 0.92[0.86-0.98];0.0116* |
| High-density lipoprotein, mmol/L | 1.11[1.01-1.21];0.0260* | 1.02[0.87-1.21];0.7681 |
| Total cholesterol, mmol/L | 0.93[0.90-0.96];<0.0001*** | 0.88[0.83-0.93];<0.0001*** |
| HbA1c, % | 0.96[0.94-0.98];0.0003*** | 1.07[1.04-1.10];<0.0001*** |
| Fasting glucose, mmol/L | 1.02[1.01-1.02];<0.0001*** | 1.03[1.02-1.04];<0.0001*** |

**Supplementary Table 4. Sensitivity analysis 1: Hazard ratios for SGLT2I v.s. DPP4I exposure effects for new onset hip fractures, and all-cause mortality in the matched cohort, with one-year lag time.**

* for p≤ 0.05, ** for p ≤ 0.01, *** for p ≤ 0.001; SGLT2I: Sodium-glucose cotransporter-2 inhibitors; DPP4I: Dipeptidyl peptidase-4 inhibitors; HR: hazard ratio; CI: confidence interval.

| **Adverse outcomes** | **SGLT2I v.s. DPP4I**  **HR [95% CI];P value** |
| --- | --- |
| All-cause mortality | 0.52[0.35-0.72];<0.0001*** |
| Hip fracture | 0.69[0.41-0.93];0.0020** |

**Supplementary Table 5. Sensitivity analysis 2: Associations of SGLT2I v.s. DPP4I on hip fractures, and all-cause mortality after propensity score matching (1:1) with competing risk models.**

* for p≤ 0.05, ** for p ≤ 0.01, *** for p ≤ 0.001; SGLT2I: Sodium-glucose cotransporter-2 inhibitors; DPP4I: Dipeptidyl peptidase-4 inhibitors; HR: hazard ratio; CI: confidence interval.

| **Models** | **Outcomes** | **SGLT2I v.s. DPP4I**  **HR [95% CI];P value** |
| --- | --- | --- |
| ***Cause-specific hazard models*** | |  |
|  | All-cause mortality | 0.58[0.41-0.62];<0.0001*** |
|  | Hip fracture | 0.66[0.45-0.90];0.0012** |
| ***Subdistribution hazard models*** | |  |
|  | All-cause mortality | 0.51[0.33-0.75];<0.0001*** |
|  | Hip fracture | 0.58[0.52-0.89];0.0145* |

**Supplementary Table 6. Sensitivity analysis 3: Hazard ratios of SGLT2I v.s. DPP4I treatment for hip fractures, and all-cause mortality in the matched cohort using different propensity matching approaches (1:1).**

* for p≤ 0.05, ** for p ≤ 0.01, *** for p ≤ 0.001; SGLT2I: Sodium-glucose cotransporter-2 inhibitors; DPP4I: Dipeptidyl peptidase-4 inhibitors; HR: hazard ratio; CI: confidence interval; PS: propensity score; IPTW: inverse probability of treatment weighting, SIPTW: stable inverse probability of treatment weighting.

| **Outcomes** | **PS stratification**  **HR [95% CI];P value** | **PS with IPTW**  **HR [95% CI];P value** | **PS with SIPTW**  **HR [95% CI];P value** |
| --- | --- | --- | --- |
| All-cause mortality | 0.52[0.41-0.66];<0.0001*** | 0.45[0.31-0.69];<0.0001*** | 0.59[0.50-0.79];<0.0001*** |
| Hip fracture | 0.62[0.51-0.88];0.0002*** | 0.68[0.42-0.97];0.0051** | 0.72[0.38-0.93];0.0011** |
